# Effect of the Healing Fit program on sleep quality, stress, and concentration: a randomized controlled study

**DOI:** 10.1101/2024.01.12.24301238

**Authors:** Wonjong Kim, Iklyul Bae, Kiyong Kim, Wonheo Ju

## Abstract

This study examined the usefulness for improving sleep quality of the Healing Fit program, which provides micro-electrical stimulation of the brain (transcranial electrical stimulation) and music therapy, in healthy adults who experienced sleep deprivation. A randomized controlled pretest–posttest design was used to evaluate effects on sleep quality, stress, and concentration. The study began after approval from the Institutional Review Board of Eulji University before conducting the study (EU22-90). Healing Fit was applied to the experimental group (n = 25) at a volume of 50 dB (about the level of normal conversation) for 30 min. Transcranial electrical stimulation intensity was set individually from 1 to 10 to the extent that the participant had no pain. Afterwards, interventions corresponding to learning, healing, and sleep music within Healing Fit were applied three times a day (30 min per session) for 14 days while participants continued their normal daily routines. The 25 control participants rested without any treatment for 30 min. Subjective/objective sleep quality, subjective/objective stress, concentration, and general characteristics were measured on day 1 of the experiment in both groups. Objective/subjective sleep quality was measured on day 7 and objective/subjective sleep quality, objective/subjective stress, and concentration were measured on day 14. The total sleep time, waking time after sleep onset, sleep efficiency, deep sleep, and subjective sleep quality were significantly better in the experimental group than in the control group. Objective and subjective stress decreased significantly in the experimental group compared to the control group, but there were no significant differences in autonomic nervous system activity. However, sympathetic and parasympathetic nervous system activity was balanced when Healing Fit was applied. There were no significant differences in concentration between groups; however, concentration tended to increase over time in the experimental group.

**Clinical Trial Registration:** Clinical Research Information Service (https://cris.nih.go.kr/; KCT0009045).

## INTRODUCTION

According to statistics from the Organisation for Economic Co-operation and Development (OECD), the average sleep time of South Koreans is 7 h 41 min, which is 41 min less than the average across the OECD countries. According to the Health Insurance Review & Assessment Service, the number of patients with insomnia in South Korea increased by 35% from 2012 to 2016. Despite increasing interest in the effect of sleep on quality of life and health among South Koreans, there are no systematic definitions and approaches to this matter [1].

Sleep accounts for one-third of the life cycle and is essential for maintaining homeostasis. Sleep affects hormone secretion, metabolic activity, the immune system, and quality of life [2,3]. Adequate sleep is essential for physical recovery and maintaining normal body functions, and positively affects psychological stability by relieving stress and tension. During sleep, the brain processes information, which can enhance memory, learning ability, and cognitive functioning [4]. Conversely, insufficient sleep causes fatigue, impairs concentration, and negatively impacts academic performance [5]. Reduced sleep leads to an increase in the prevalence of diseases such as insomnia, and a rise in social costs, such as those caused by drowsiness-induced traffic accidents, the worsening of chronic conditions, and depression diagnosis and treatment [1]. Furthermore, sleep is related to interactions within the autonomic nervous system (ANS), with proper sleep increasing the activity of the parasympathetic nervous system (PNS), lowering body temperature, stabilizing the heart rate and breathing, and accelerating recovery. However, if sympathetic nervous system (SNS) activity increases, metabolism becomes more active and affects the heart rate, blood pressure, and respiration, making it difficult to fall asleep or enter deep sleep with resultant poor sleep quality and daily stress [6,7].

Stress refers to the tension felt in a situation where adaptation to the environment does not function well. When exposed to stress, the SNS is activated, speeding up the body’s metabolism. Once the SNS is activated, blood pressure and heart rate increase, and the hypothalamic–pituitary–adrenal axis is activated, increasing the secretion of cortisol, a representative stress hormone [8]. Such metabolic acceleration disrupts the balance of the ANS, making it difficult to fall asleep and enter deep sleep. If this situation persists, proper physical and psychological recovery is not achieved, resulting in fatigue and impaired concentration [4].

Concentration is the ability to focus effort and attention on a specific task. Self-control is essential to maintain concentration, and a lack of self-control can make it impossible to sustain that focus. By maintaining a high level of concentration, individuals can not only increase the efficiency of their learning but also achieve their goals more easily across various activities [9].

Healing Fit is a device that applies transcranial electrical stimulation (tES) and sound therapy. tES is a non-invasive and painless procedure that stimulates targeted cortical areas by applying a micro-electric current to two electrodes placed on the skull [10,11]. The microcurrent flowing between the two electrodes induces changes in neural excitability and activity through specific molecular mechanisms that mediate synaptic plasticity [12]. A literature review of tES-related studies confirmed positive effects, such as the enhancement of cognitive functions, including memory and learning, and improved subjective sleep quality [13–15].

While tES-related studies have shown its positive impact on subjective sleep measures, the findings related to objective sleep quality are not clear, indicating the need for scientific evidence through further research. In this context, this study was conducted to examine the effect of Healing Fit on objective and subjective sleep parameters, stress, balance of the ANS, and concentration in adults with impaired sleep quality and to provide foundational data for future research.

## MATERIALS AND METHODS

### Study design

This study adopted a randomized controlled group pretest–posttest design to apply the Healing Fit program and assess its effects on sleep quality, stress, and concentration.

### Participants

The participants were adults residing in the Seoul, Uijeongbu, Daejeon, and Gimcheon regions of South Korea who had experienced sleep deprivation. The target population was healthy adults aged between 20 and 60 years, recruited through a public announcement. Participants gave prior written consent at the time of recruitment, and it was explained to them that the study would be conducted from February 27, 2023 to April 2, 2023.

The inclusion criteria were: 1) consented to participate in the study after understanding its purpose and objectives; 2) being conscious and able to communicate; 3) 20–60 years of age; 4) both men and women; and 5) no diseases affecting hearing. The exclusion criteria were: 1) on medication for physical or mental illnesses (e.g., anti-anxiety drugs, sleeping pills, painkillers), and 2) affected by chronic disease. The study was conducted with volunteers who applied after the recruitment announcement. The study was approved by the Institutional Review Board of Eulji University in Uijeongbu (approval number: EU22-90).

### Sample size calculation

The minimum sample size was determined using G*power 3.1.9 by inputting a significance level of 0.05, power (1-β) of 0.80, and effect size of 0.25. The medium effect size of 0.25 was derived from Cohen’s f. As a result, a total required sample size of 44 was calculated, and 50 participants were recruited, taking a dropout rate of 10% into account.

### Participant allocation and blinding

Fifty participants were randomized into experimental and control groups (25 each) using block blind randomization based on random number generation in Microsoft Excel (Microsoft, Redmond, WA). All data from the 50 participants were collected and used for analysis without any dropouts during the data collection process (Fig 1).

**Fig 1.**
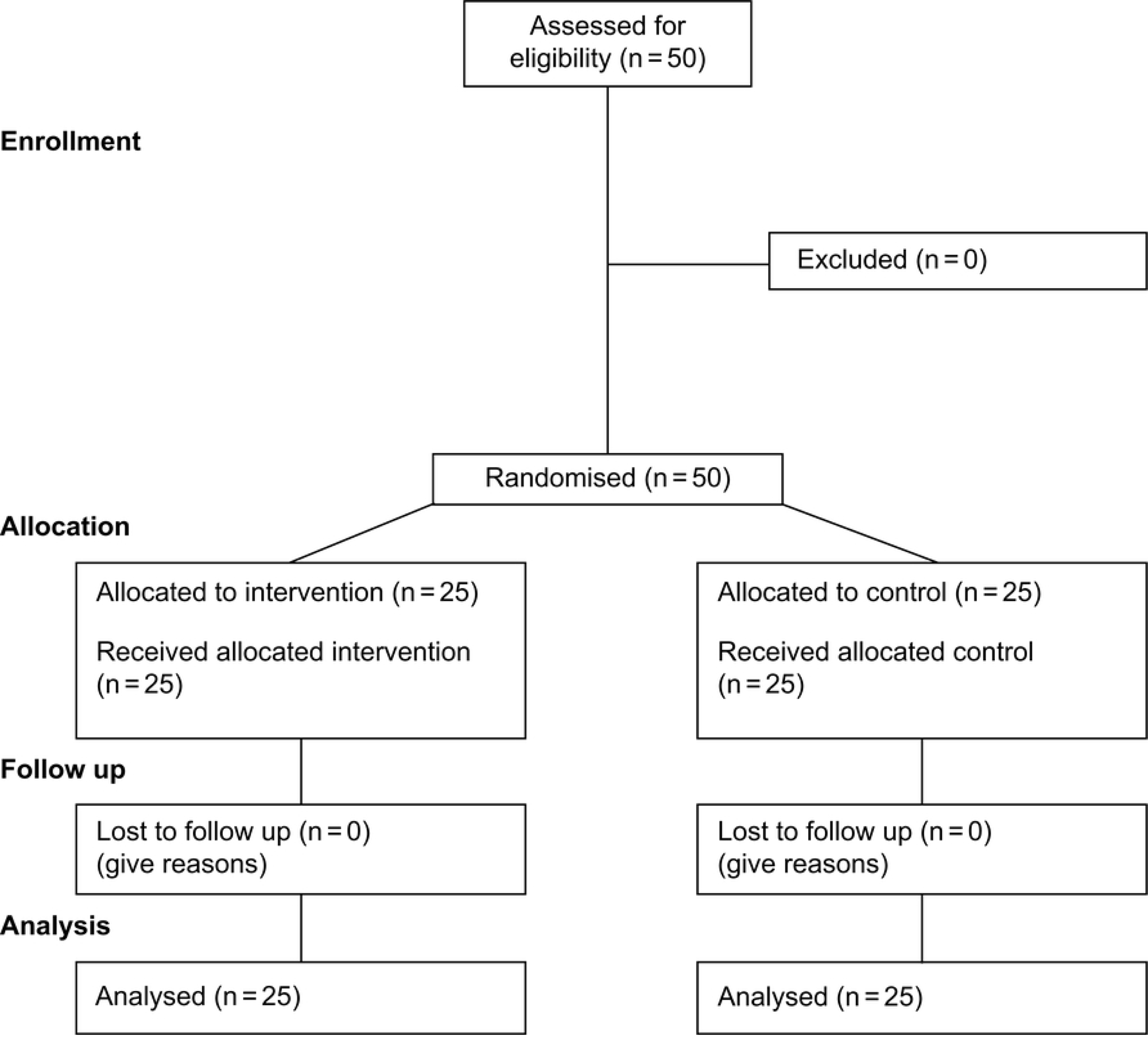
CONSORT flow diagram.

Furthermore, to minimize the diffusion effect, data were collected from the control group first, followed by the experimental group’s treatment and data collection upon completion of the treatment. One research assistant carried out both the pretest and posttest, and researchers were blinded to the allocation to enhance the validity of the study.

### Experimental treatment

The laboratory occupies an area of 19.83 m^2^, and the indoor temperature was set at 25°C, taking into consideration the optimal conditions for measuring stress levels, heart rate, blood pressure, autonomic balance, and concentration. The laboratory featured windows to ensure proper ventilation and comfortable furniture, such as sofas, tables, and chairs, to create a welcoming environment for the participants. Moreover, specialized equipment for monitoring stress, heart rate, autonomic balance, and measuring blood pressure was installed.

Healing Fit is a device that provides tES and sound therapy. tES sends a weak electrical signal to the surface of the brain through electrodes located on the scalp, inducing spontaneous nerve cell activity, normalizing brain function, and increasing the secretion of endorphins, thereby managing stress and controlling pain. Concurrently, sound therapy delivers a frequency of 200 Hz to the left ear and 210 Hz to the right ear through stereo headphones. The brain perceives this sound as the difference between the two frequencies (i.e., 10 Hz), referred to as the third signal sound or binaural beat. This signal resonates with the brain’s neural nucleus and sends the signal to the cerebral cortex, altering existing brain waves. Sound therapy consists of healing, sleep, and learning modes. The effects of sound therapy include reducing cortisol secretion, increasing natural killer cells, and restoring balance in the ANS.

The experimental group was treated with Healing Fit for 30 min at a volume of 50 dB, equivalent to the sound level of normal conversation. tES intensity was individually adjusted within a range of 1 to 10, where the participant did not experience pain. Participants were introduced to the operation of Healing Fit and its application was demonstrated in the laboratory. Initially applied at level 5, subsequent adjustments were made according to the presence or absence of pain, and the applicable intensity levels were determined and taught to the participants. The experimental group was instructed to apply the music interventions for learning, healing, and sleep within the Healing Fit program three times a day (30 min per session) for 14 days while engaging in daily activities. They were informed that if pain was felt during application, they could reduce the intensity to 1, and if the application became difficult due to pain, they could stop the experiment at any time. The control group did not receive any treatment and continued their daily activities.

### Research tools

Objective sleep quality was determined by measuring the following values using FitBit Charge 4 (FB407; FitBit Inc., San Francisco, CA):

① Total sleep time: the total time (min) that the participant slept.
② Waking after sleep onset: the total time spent awake (min) between falling asleep and waking up.
③ Deep sleep: Stage 3 non-rapid-eye-movement sleep (min) measured as deep sleep on FitBit Charge 4.
④ Sleep efficiency: percentage (%) of total sleep time excluding the time awake after sleep onset

Sleep quality was assessed using the Korean Modified Leeds Sleep Evaluation Questionnaire (KMLSEQ) [16]. The KMLSEQ consists of 10 items, each measured on a 100-mm visual analog scale (VAS), evaluating the ease of falling asleep, the quality of sleep, the ease of awakening, and behavior following waking. The sleep score is determined by dividing the sum of all item scores by the number of items, resulting in a range from 0–100. Higher scores indicate better sleep quality, and a score of 66 is considered a cutoff point, with scores below this threshold deemed to represent poor sleep quality. Objective stress was assessed through a stress index, calculated by quantifying the balance of the ANS (hereinafter referred to as “ANS balance”) using a 12-lead echocardiogram. This calculation was based on heart rate variability (HRV) data, continuously measured for a duration of 5 min using the Canopy9 RSA system (IEMBIO, Chuncheon, Korea). The stress index has a range of 1–10, with higher values reflecting a greater degree of exposure to stress.

Subjective stress was measured with the VAS, in which the participant indicates the perceived level of stress on a 10-cm horizontal line marked with numbers from 0–10, with higher scores meaning higher perceived stress.

ANS balance is the interaction between the parasympathetic and sympathetic components of the ANS. It can be assessed by continuously measuring HRV for 5 min and measuring the low-frequency (LF) and high-frequency (HF) bands of HRV, which are associated with the SNS and PNS, respectively. The higher the SNS activity (LF) and the lower the PNS activity (HF), the higher the stress level, with a higher ANS balance (LF/HF ratio) indicating more stress.

Concentration levels were assessed using Part B of the Trail Making Test, included in Reitan’s neuropsychological test battery [17]. This test, primarily used to evaluate cognitive function with a focus on concentration, requires the test-taker to use a writing tool to draw lines connecting elements in the order of 1-A-2-B-3-C, with the time taken to complete the task being measured. If the test-taker accidentally lifts their hand off the paper, an immediate correction is made, and the final time taken to complete the task without mistakes is recorded.

### Data Collection

Data were collected from the control group from February 27, 2023 to April 2, 2023, and from the experimental group from April 3, 2023 to May 7, 2023, after administration of the Healing Fit program as explained above.

Before administration of the Healing Fit program to the experimental group, general characteristics, subjective sleep quality, subjective and objective stress, ANS balance (LF/HF ratio), heart rate, blood pressure, and concentration level were measured. After baseline measurements (pretest), the experimental group received Healing Fit training in the laboratory. During Healing Fit application, the intensity of tES was set at a level where the participant felt no pain. Prior to initial application, the best intensity for each participant was tested by varying the intensity among five levels. Additionally, they were trained to wear the Fitbit Charge 4 while sleeping. The experimental group followed the Healing Fit program’s learning, healing, and sleep music interventions three times a day (30 min per session) for 14 days while continuing their routine daily lives. The baseline measurement of objective sleep quality refers to the results of sleep patterns transmitted to a smartphone from the Fitbit Charge 4 worn on the day they received training in the laboratory. From the day after the first sleep, the experimental group applied the Healing Fit device three times a day. On day 7, the experimental group sent the sleep quality results to the research assistant via smartphone, and on day 14, post-intervention measurements were taken for objective and subjective sleep quality, objective and subjective stress, ANS balance, and concentration.

The control group underwent the same measurements as the experimental group at baseline: general characteristics, subjective sleep quality, subjective and objective stress, ANS balance (LF/HF ratio), heart rate, and concentration. The control group was also trained to wear the Fitbit Charge 4 while sleeping. They then spent 14 days living their routine daily lives without intervention and sent the results of sleep quality to the research assistant via smartphone on day 7. On day 14, post-measurements were taken for objective and subjective sleep quality, objective and subjective stress, ANS balance, and concentration.

### Data Analysis

The collected data was analyzed using SPSS for Windows version 26.0 (IBM, Armonk, NY). The homogeneity of the general characteristics of the participants was analyzed using frequency, percentage, mean, *t*-test, and the χ^2^-test. The homogeneity of the dependent variables was analyzed using the *t*-test. *T*-tests and repeated-measures analysis of variance were used to evaluate the treatment effects between the experimental and control groups. Additionally, the partial η^2^ was analyzed to explain the effect size between independent and dependent variables. The effect size was small, medium, or large if the partial η^2^ was 0.01, 0.06, or 0.14, respectively. Therefore, the closer the value of partial η^2^ was to 1, the greater the mean difference between the groups and the smaller the error. The *P*-value for hypothesis testing was set at 0.05.

After completing data collection with the experimental group, the control group received the same intervention. All members of the control group were administered the Healing Fit program for 14 days in the same manner as the experimental group, except for those who opted out of participating in the experimental intervention.

## RESULTS

### Baseline homogeneity between the experimental and control groups

The general characteristics of the 50 participants who took part in the study are outlined in Table 1. Participants were randomly assigned to either the Healing Fit non-application group (control group) or the Healing Fit application group (experimental group), with 25 in each group. The control group included 11 males (44%) and 14 females (56%) and the experimental group included 8 males (32%) and 17 females (68%). The mean ages and subjective sleep times of the control and experimental groups were 28.88 ± 4.64 and 27.64 ± 5.44 years, respectively, and 405.60 ± 72.00 and 406.80 ± 75.54 min, respectively. No intergroup differences were observed in sex, age, subjective sleep time, marital status, education level, occupation type, number of family members, smoking status, or drinking habits.

**Table 1.** Participants’ general characteristics and the results of homogeneity testing of dependent variables (n = 50)

| Variables | Categories | Cont. G | Exp. G | $X^2$ or $t$ | p |
| --- | --- | --- | --- | --- | --- |
|  |  | (n = 25) | (n = 25) |  |  |
| | | Mean ± SD or N<br>(%) | Mean ± SD or N<br>(%) | | |
| Sex | Male | 11(44.0) | 8(32.0) | -0.863 | 0.392 |
|  | Female | 14(56.0) | 17(68.0) |  |  |
| | Age (years) | 28.88 ± 4.64 | 27.64 ± 5.44 | 0.226 | 0.822 |
| | Subjective sleep time (min) | 405.60 ± 72.0 | 406.80 ± 75.54 | -0.057 | 0.954 |
| Marital status | Married | 9(36.0) | 8(32.0) | -0.335 | 0.793 |
|  | Unmarried | 16(64.0) | 17(68.0) |  |  |
| Education | High school | 9(36.0) | 8(32.0) | -0.257 | 0.798 |
|  | Undergraduate | 15(60.0) | 16(64.0) |  |  |
|  | Postgraduate or higher | 1(4.0) | 1(4.0) |  |  |
| Occupation | Office | 6(24.0) | 6(24.0) | -0.592 | 0.557 |
|  | Professional | 13(52.0) | 12(48.0) |  |  |
|  | Self-employed | 6(24.0) | 7(28.0) |  |  |
| Smoking | Yes | 15(60.0) | 12(48.0) | -0.840 | 0.405 |
|  | No | 10(40.0) | 13(52.0) |  |  |
| Drinking | Yes | 22(88.0) | 20(80.0) | -0.760 | 0.451 |
|  | No | 3(12.0) | 5(20.0) |  |  |
| | Subjective sleep quality | 47.64 ± 9.15 | 48.56 ± 8.96 | -0.359 | 0.721 |
| | Subjective stress (VAS) | 5.48 ± 0.96 | 5.44 ± 0.96 | 0.147 | 0.884 |
| Objective sleep quality | Total sleep time (min) | 404.84 ± 29.12 | 392.48 ± 26.46 | 1.571 | 0.123 |
| | Waking after sleep onset (min) | 76.40 ± 7.95 | 76.56 ± 6.46 | -0.078 | 0.938 |
| | Sleep efficiency (%) | 81.07 ± 2.01 | 80.49 ± 1.08 | 1.289 | 0.203 |
|  | Deep sleep (%) | 16.00 ± 2.08 | 15.84 ± 1.84 | 0.288 | 0.755 |
| Objective stress | Stress index | 6.44 ± 0.92 | 6.16 ± 0.85 | 1.120 | 0.268 |
|  | High-frequency activity | 4.38 ± 0.72 | 4.20 ± 0.60 | 0.967 | 0.339 |
|  | Low-frequency activity | 5.90 ± 0.81 | 6.09 ± 0.78 | -0.803 | 0.426 |
| Vital signs | Heart rate (bpm) | 74.88 ± 7.58 | 75.04 ± 8.56 | -0.070 | 0.944 |
|  | SBP (mmHg) | 114.96 ± 8.86 | 112.80 ± 8.74 | 0.868 | 0.390 |
|  | DBP (mmHg) | 76.24 ± 7.25 | 72.48 ± 7.67 | 1.781 | 0.081 |
|  | Concentration | 83.92 ± 21.34 | 84.64 ± 13.02 | -0.144 | 0.886 |

Exp. G: experimental group; Cont. G: control group; SD: standard deviation; VAS: visual analog scale; SBP:
systolic blood pressure; DBP: diastolic blood pressure. \*p < 0.05

Table 1 also presents the results of homogeneity testing of the dependent variables: stress, sleep quality, and concentration. The subjective sleep quality score was 47.64 in the control group and 48.56 in the experimental group. Subjective stress levels were 5.48 in the control group and 5.44 in the experimental group. A comparison of the variables associated with objective sleep quality between groups yielded the following results: total sleep time was 404.84 min for the control group vs. 392.48 min for the experimental group, waking after sleep onset was 76.40 vs. 76.56 min, sleep efficiency was 81.07% vs. 80.49%, and deep sleep duration was 16.00 vs. 15.84 min. No significant differences were observed between groups in terms of subjective sleep quality or subjective stress.

A comparison of objective stress between the control and experimental groups yielded the following results: the objective stress index was 6.44 vs. 6.16, with HF activity of 4.38 vs. 4.20 and LF activity of 5.90 vs. 6.09; among the vital signs, heart rate was 74.88 vs. 75.04 bpm, systolic blood pressure was 114.96 vs. 112.80 mmHg, and diastolic blood pressure was 76.24 vs. 72.48 mmHg; and concentration level was 83.92% vs. 84.64%, respectively. No significant differences were observed between groups in terms of objective stress, vital signs, or concentration.

### Effect of Healing Fit on sleep quality, stress, and concentration

Objective sleep quality was measured three times: at baseline, on day 7, and on day 14. The mean total sleep time measured on day 7 was significantly different between the control and experimental groups, with 392.92 vs. 407.16 min (p = 0.017), as did the mean total sleep time measured on day 14 (391.88 vs. 411.68 min, p = 0.023). Repeated measurements of total sleep time over 14 days showed no significant difference (p = 0.175) based on time or group but showed a significant interaction effect between group and time (p < 0.001). The effect size (η²) of the treatment based on group and time was 0.130 (Table 2).

**Table 2.**
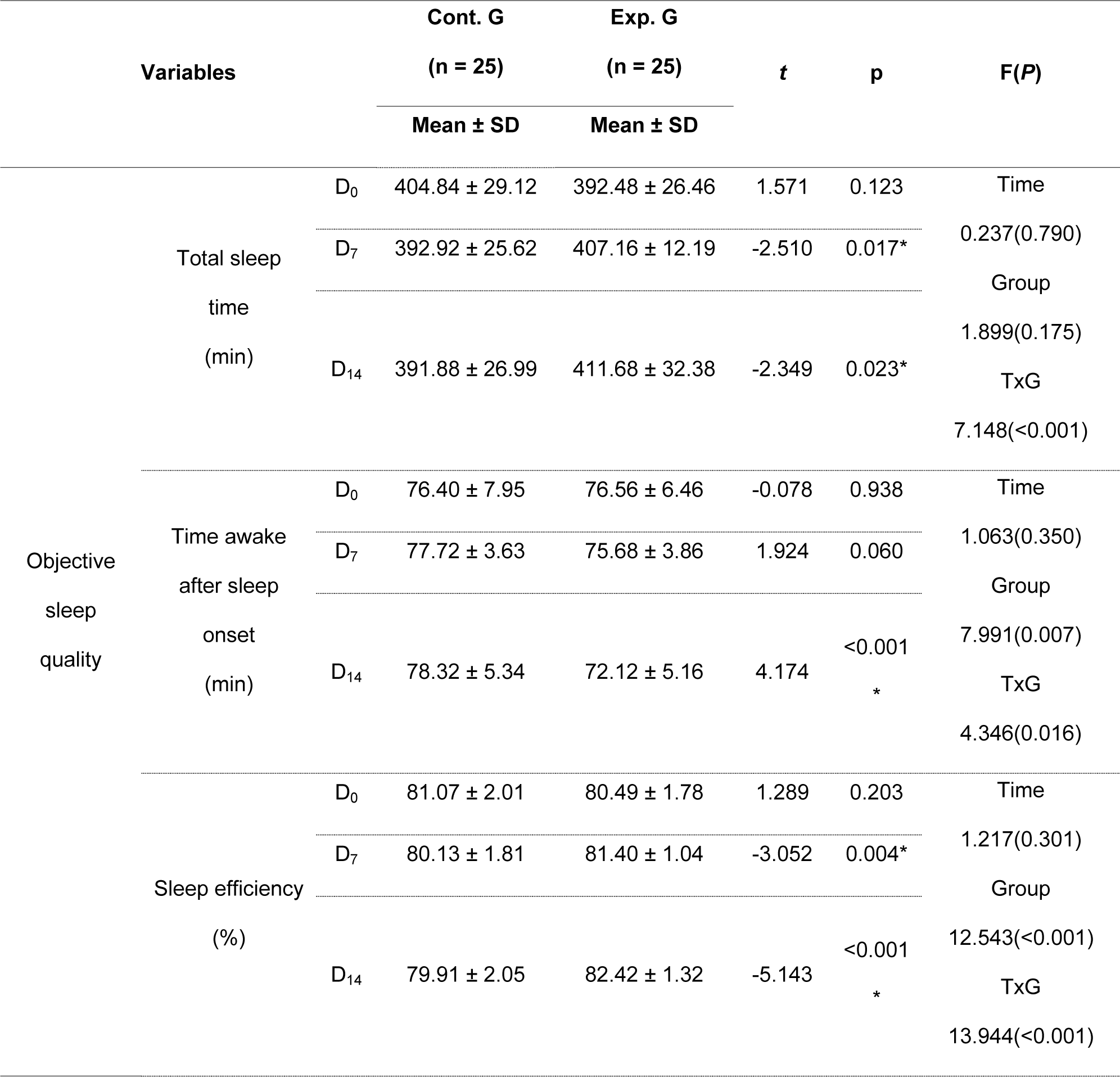
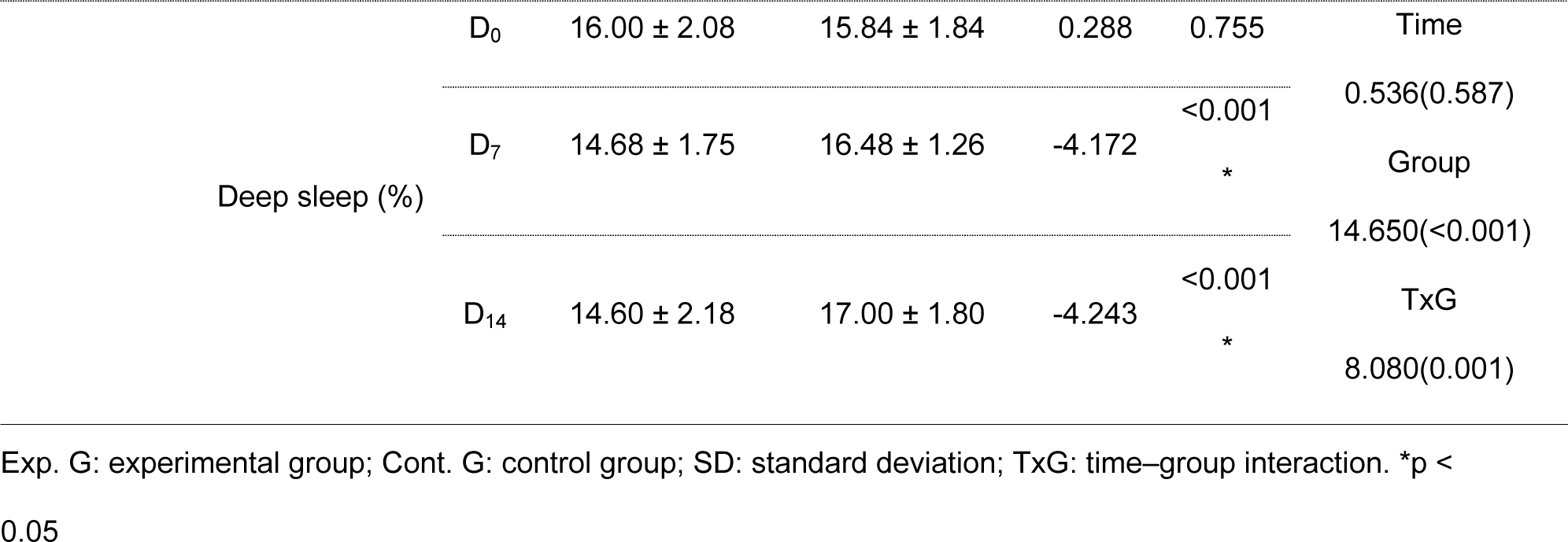
Comparison of objective sleep quality between the experimental and control groups.

Mean waking after sleep onset measured on day 7 was not significantly different between the control and experimental groups (77.72 and 75.68 min, respectively, p = 0.060). However, a significant intergroup difference was shown on day 14 (78.32 vs. 72.12 min, respectively, p < 0.001). Repeated measurements over 14 days showed no significant effect of time (p = 0.350) but a significant effect of group (p = 0.007) and a significant interaction effect between group and time (p = 0.016). The effect size (η²) of the treatment based on group and time was 0.083 (Table 2, Fig 2).

**Fig 2.**
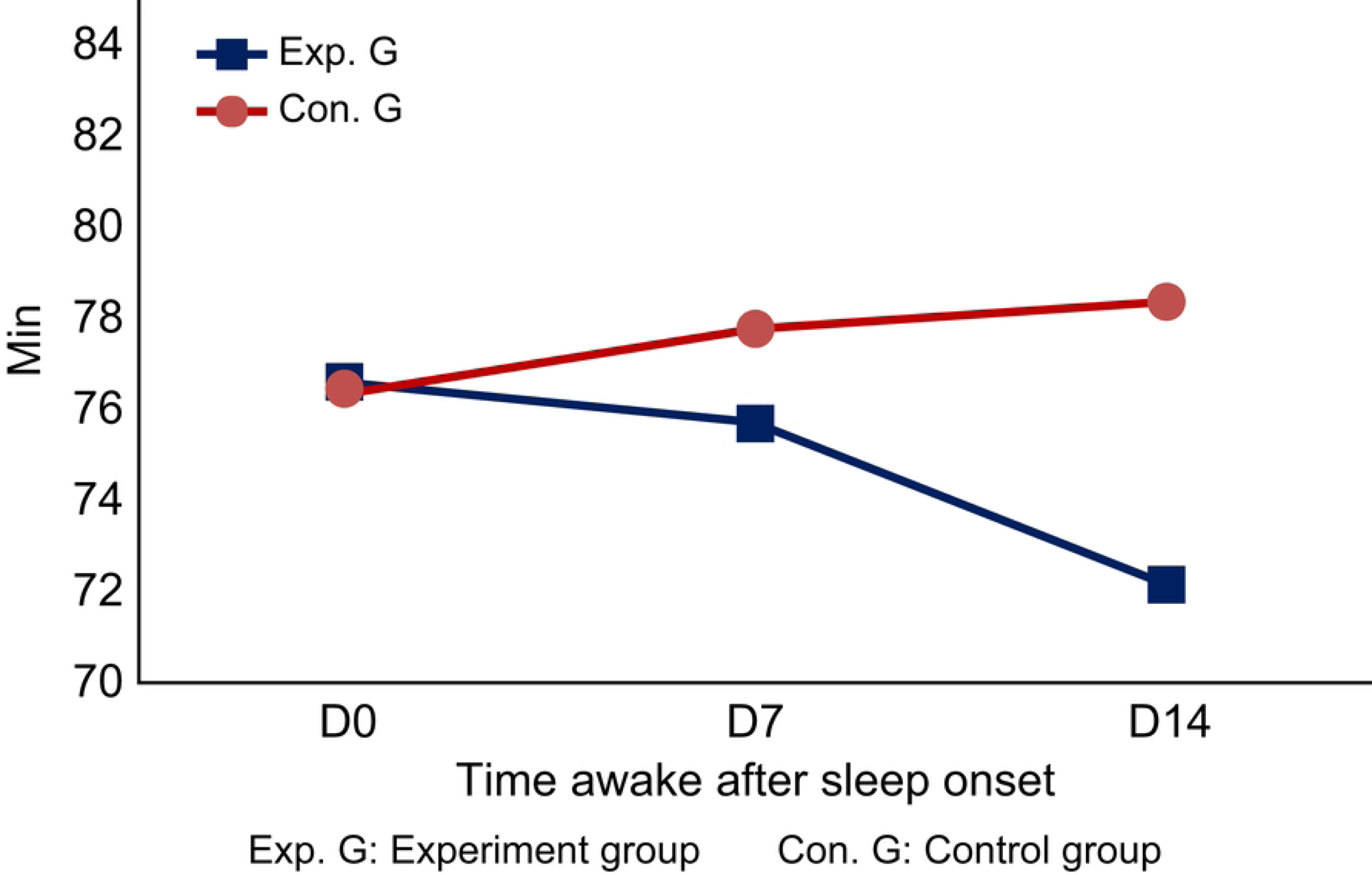
Comparison of waking after sleep onset (min) between the experimental and control groups. D, day.

Sleep efficiency was significantly different between the control and experimental groups at day 7 (80.13% vs. 81.40%, respectively, p = 0.004) and day 14 (79.91% vs. 82.42%, respectively, p < 0.001). Repeated measurements over 14 days showed no significant effect of time (p = 0.301) but a significant effect of group (p < 0.001) and a significant interaction effect between group and time (p < 0.001). The effect size (η²) of the treatment based on group and time was 0.225 (Table 2, Fig 3).

**Fig 3.**
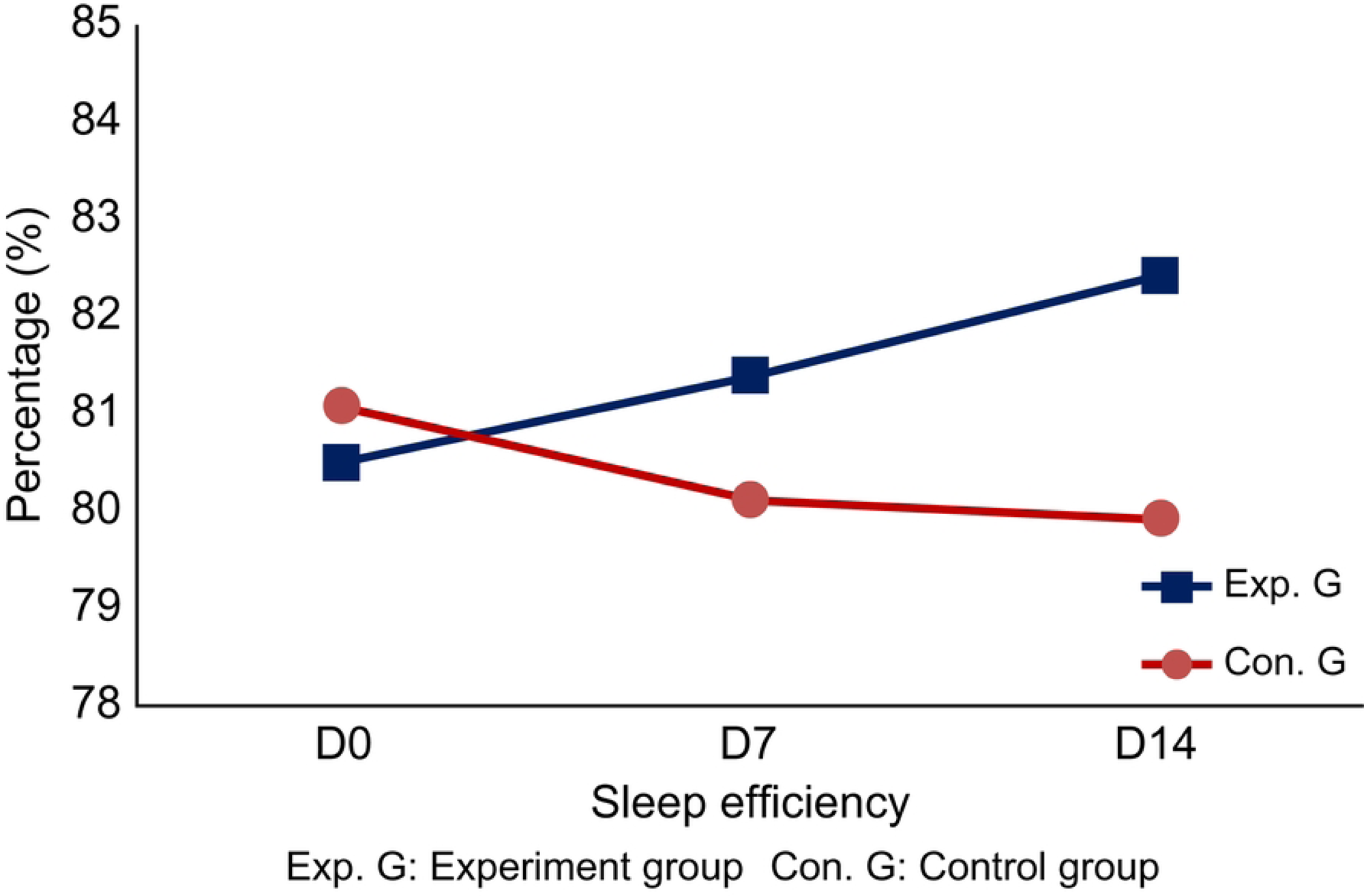
Comparison of sleep efficacy (%) between the experimental and control groups. D, day.

Deep sleep (%) was significantly different between the control and experimental groups on day 7 (14.68% vs. 6.48%, respectively, p < 0.001) and day 14 (14.60% vs. 17.00%, respectively, p < 0.001). Repeated measurements of deep sleep over 14 days showed no significant effect of time (p = 0.587) but a significant effect of group (p < 0.001), as well as an interaction effect between group and time (p < 0.001). The effect size (η²) of the treatment based on group and time was 0.144 (Table 2, Fig 4).

**Fig 4.**
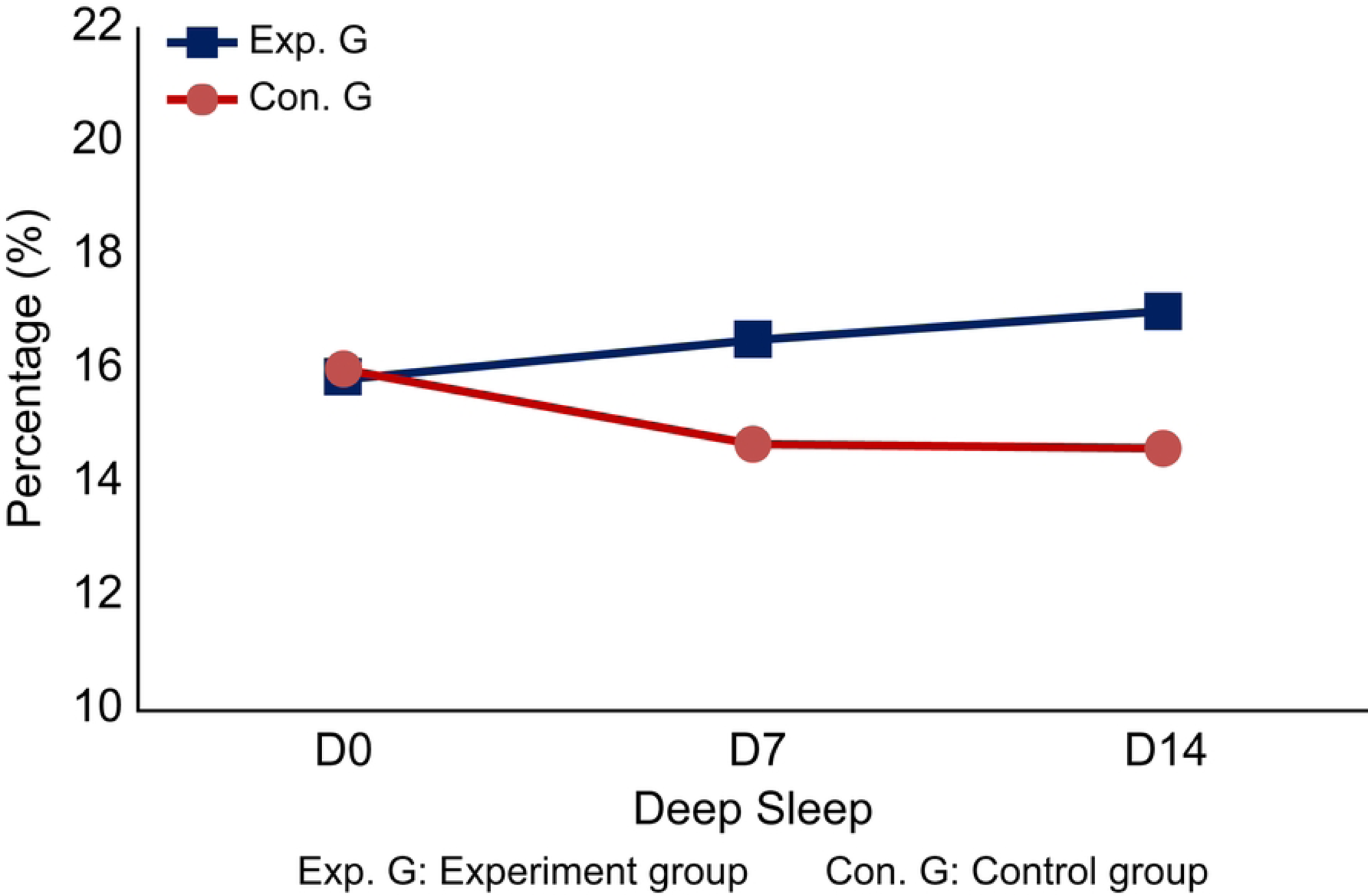
Comparison of deep sleep (%) between the experimental and control groups. D, day.

Subjective sleep quality measurements are presented in Table 3. At baseline, the mean subjective sleep quality scores were 47.64 for the control group and 48.56 for the experimental group. On day 14, the mean scores differed significantly, with the control group scoring 49.88 and the experimental group scoring 56.52 (p = 0.009).

**Table 3.** Comparison of subjective sleep quality between the experimental and control groups.

| Variables |  | Cont. G<br>(n = 25) | Exp. G<br>(n = 25) | t | p |
| --- | --- | --- | --- | --- | --- |
| | | Mean ± SD | Mean ± SD | | |
| Subjective sleep quality | D<sub>0</sub> | 47.64 ± 9.15 | 48.56 ± 8.96 | -0.359 | 0.721 |
| | D<sub>14</sub> | 49.88 ± 8.55 | 56.52 ± 8.75 | -2.715 | 0.009* |

Exp. G: experimental group; Cont. G: control group; SD: standard deviation. \* $p < 0.05$

Table 4 presents the objective stress measurement results. At baseline, the mean stress index was 6.44 for the control group and 6.16 for the experimental group. On day 14, the mean scores were 6.16 for the control group and 3.52 for the experimental group, showing a significant difference between the two groups (p < 0.001) (Table 4).

**Table 4.**
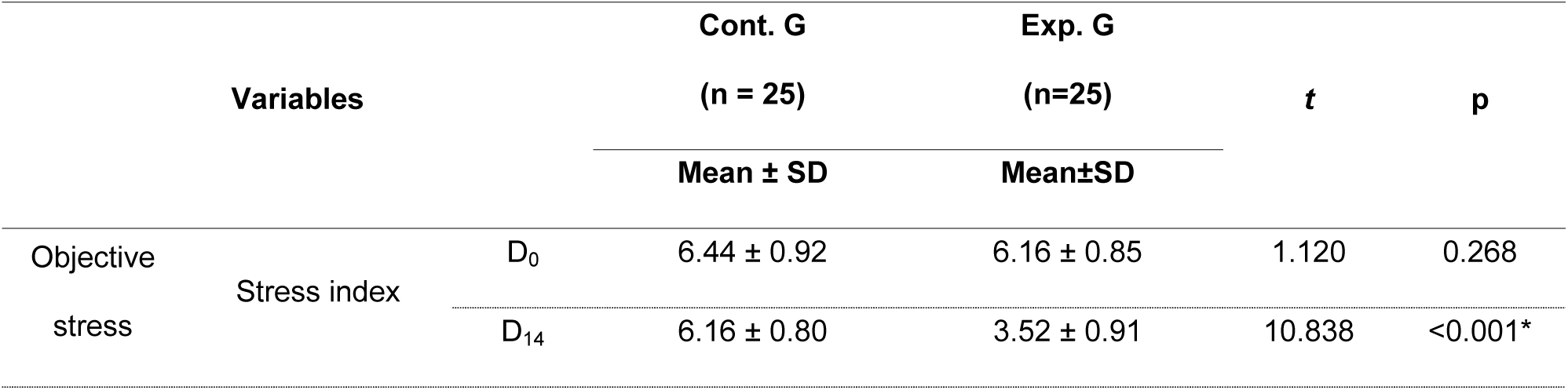
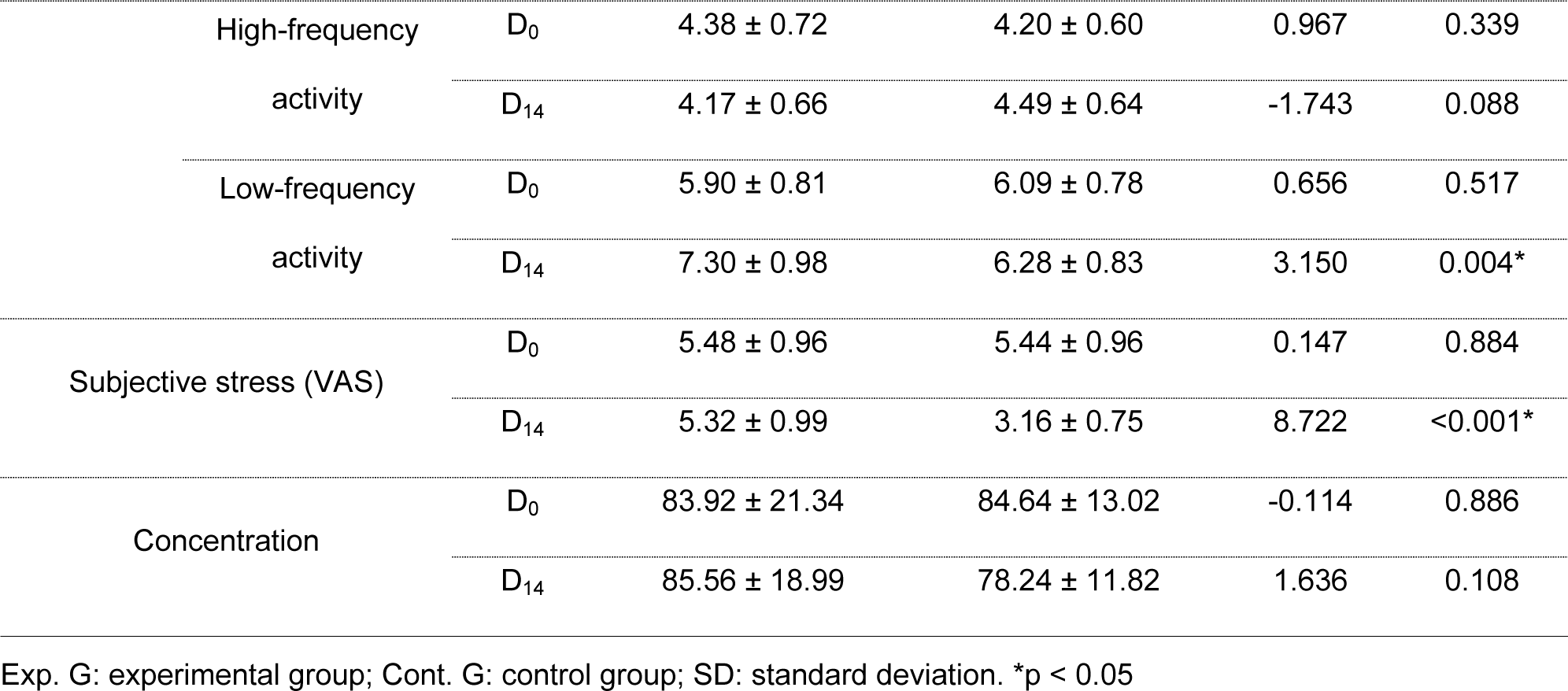
Comparison of subjective and objective stress between the experimental and control groups.

| Variables |  |  | Cont. G<br>(n = 25) | Exp. G<br>(n=25) | t | p |
| --- | --- | --- | --- | --- | --- | --- |
| | | | Mean ± SD | Mean ± SD | | |
| Objective stress | Stress index | D<sub>0</sub> | 6.44 ± 0.92 | 6.16 ± 0.85 | 1.120 | 0.268 |
| | | D<sub>14</sub> | 6.16 ± 0.80 | 3.52 ± 0.91 | 10.838 | <0.001* |

|  |  |  |  |  |  |
| --- | --- | --- | --- | --- | --- |
| High-frequency activity | D<sub>0</sub> | 4.38 ± 0.72 | 4.20 ± 0.60 | 0.967 | 0.339 |
|  | D<sub>14</sub> | 4.17 ± 0.66 | 4.49 ± 0.64 | -1.743 | 0.088 |
| Low-frequency activity | D<sub>0</sub> | 5.90 ± 0.81 | 6.09 ± 0.78 | 0.656 | 0.517 |
|  | D<sub>14</sub> | 7.30 ± 0.98 | 6.28 ± 0.83 | 3.150 | 0.004* |
| Subjective stress (VAS) | D<sub>0</sub> | 5.48 ± 0.96 | 5.44 ± 0.96 | 0.147 | 0.884 |
|  | D<sub>14</sub> | 5.32 ± 0.99 | 3.16 ± 0.75 | 8.722 | <0.001* |
| Concentration | D<sub>0</sub> | 83.92 ± 21.34 | 84.64 ± 13.02 | -0.114 | 0.886 |
|  | D<sub>14</sub> | 85.56 ± 18.99 | 78.24 ± 11.82 | 1.636 | 0.108 |

Exp. G: experimental group; Cont. G: control group; SD: standard deviation. \*p < 0.05

The mean HF activity results at baseline were 4.38 for the control group and 4.20 for the experimental group. On day 14, the mean scores were 4.17 for the control group and 4.49 for the experimental group, showing no significant difference between groups (Table 4).

The mean LF activity results at baseline were 5.90 for the control group and 6.09 for the experimental group. On day 14, the mean scores were 7.30 for the control group and 6.28 for the experimental group, showing a significant difference between groups (p = 0.004) (Table 4).

Subjective stress measurements are presented in Table 4. At baseline, the mean subjective stress score was 5.48 for the control group and 5.44 for the experimental group. On day 14, the mean scores differed significantly, with the control group scoring 5.32 and the experimental group scoring 3.16 (p < 0.001).

Concentration measurements are presented in Table 4. At baseline, the mean concentration durations were 83.92 s for the control group and 84.64 s for the experimental group. On day 14, no significant difference was observed between groups (85.56 vs. 78.24 s, p < 0.001) (Table 4).

## DISCUSSION

This study aimed to evaluate an intervention to enhance sleep quality and concentration and alleviate stress in adults with compromised sleep health. To achieve this, we introduced the Healing Fit program utilizing tES. Over a span of 2 weeks, the experimental group subjected to Healing Fit exhibited a significant enhancement in both objective and subjective sleep quality when compared to the control group.

Objective sleep quality was analyzed based on metrics from the FitBit Charge 4 and encompassed total sleep time, waking after sleep onset, deep sleep, and sleep efficiency. Total sleep time, sleep efficiency, and the duration of deep sleep increased with Healing Fit, while the frequency of waking after sleep onset decreased, indicating an overall improvement in sleep quality. A prior study [18] also found that tES had a positive effect on objective sleep quality. Although different tools were used to measure objective sleep quality, both studies found an increase in objective sleep quality. In comparison with the previous study, our study involved a longer intervention (14 days vs. 3 sessions) and a larger sample size (50 vs. 13). Furthermore, the accuracy of the results of this study was established through a convenient intervention application.

Similarly, over the course of 2 weeks, there was a significant increase in the subjective sleep quality score of the experimental group compared to the control group. As reported in a prior study [4], tES positively impacted perceived improvement in subjective sleep quality. Our findings are consistent with another study [19] that suggested that psychological comfort associated with the treatment contributed to enhanced sleep quality and stress alleviation.

The overall results of the Healing Fit intervention indicate significant improvements in both objective and subjective sleep quality. The duration and proportion of deep sleep increased over time in the experimental group. This enhancement in objective sleep quality is likely to have influenced the perceived improvement in subjective sleep quality.

Following the 2-week Healing Fit intervention, the experimental group demonstrated a significant reduction in both objective and subjective stress compared to the control group. Stress can arise from external triggers but can also be due to accumulated physical fatigue and decreased resilience. It is imperative to manage stress in everyday life, given its association with cardiovascular diseases such as stroke, as well as various other chronic conditions [20]. Among the methods available for alleviating stress and aiding physical recovery, sleep stands out as particularly pivotal. Enhanced sleep quality exerts a positive influence on physical recuperation and is a key factor in stress reduction [21].

Objective stress was assessed using an ANS measuring device (Canopy 9) and subjective stress was assessed using a VAS. Following a 2-week intervention with Healing Fit, the experimental group exhibited a notable decrease in both objective and subjective stress compared to the control group. As indicated by an earlier study [22], tES was found to be effective in activating endorphins and reducing objective stress, which was in line with our findings. The decrease in both objective and subjective stress presumably contributed to improved sleep quality. Based on these favorable outcomes for subjective and objective sleep quality, it can be deduced that the application of Healing Fit resulted in increased deep sleep time and augmented sleep efficiency. This likely contributed to enhanced physical recovery and relaxation, further facilitating stress alleviation.

Over the 2-week Healing Fit intervention, the ANS balance measurements in the experimental group did not show significant differences when compared to the control group. Nevertheless, a prior study [23] found that tES reduced SNS activity while augmenting PNS activity. Although our study echoed these findings in terms of a positive shift in the balance between the SNS and PNS, it did not reach statistical significance. The LF/HF ratio serves as a significant marker for autonomic balance. An ideal LF/HF ratio is commonly understood to be 1.5 [24]. This ratio is considered more telling than standalone numerical statistics. In our study, the LF/HF ratio in the experimental group was close to 1.5. This implies that the Healing Fit application, through the provision of tES, mitigated stress stemming from physical fatigue and stabilized physiological indicators through enhanced sleep quality.

The concentration level measured over the 2-week Healing Fit intervention showed no significant difference between the experimental and control groups. In a prior study [25], however, tES had a positive effect on cognitive abilities related to memory and learning. This discrepancy might have arisen from differences in the participants’ characteristics and measurement durations between the two studies. However, a pretest–posttest comparison did reveal an increase in the concentration level of the experimental group compared to the control group. It is hypothesized that Healing Fit influenced physical stability, stress levels, relaxation, and cognitive ability. In future research, extending the duration of Healing Fit application may yield more accurate and meaningful findings related to concentration.

Overall, Healing Fit showed significant enhancements in both objective and subjective sleep qualities. This signifies that the use of Healing Fit not only boosts objective sleep quality and sleep efficiency but also positively impacts perceived sleep quality. Superior sleep quality facilitates the maintenance of homeostasis, harmonizes the balance between the SNS and PNS, and ensures stability in vital signs, such as blood pressure and heart rate. Moreover, enhanced sleep quality can reduce stress, bolster psychological stability, and augment cognitive abilities such as memory and learning.

This study is significant as it introduces the Healing Fit intervention as a possible treatment for adults struggling with sleep duration and quality. Healing Fit presents a more user-friendly approach with fewer side effects than conventional pharmacological treatments. Furthermore, Healing Fit’s positive impact extends beyond sleep to aspects, such as stress and ANS balance. Its compact design offers the advantage of easy portability, enabling usage at any time or place. Based on these results, it is expected that applying the Healing Fit program to individuals experiencing sleep difficulties could enhance their quality of life.

### Conclusions and Limitations

This study verified the efficacy of the Healing Fit application by demonstrating significant outcomes in objective and subjective sleep quality as well as stress. Healing Fit not only improved objective and subjective sleep quality and sleep efficiency but also aided in maintaining homeostasis. It ensured an appropriate balance between the SNS and PNS and positively influenced cognitive abilities. The efficacy of Healing Fit was established in this study by providing adults experiencing sleep difficulties and impaired sleep quality with an intervention that has fewer side effects that is easier to apply than pharmacological treatments. The results of this study highlight the feasibility of Healing Fit as an intervention beneficial for sleep and stress management. It is anticipated that applying it to a broader group of adults struggling with sleep issues could significantly enhance their quality of life.

This study recruited adults experiencing impaired sleep quality from the Seoul, Uijeongbu, Daejeon, and Gimcheon regions of South Korea. Given the regional variations in living environments and occupational backgrounds, it is crucial to approach the generalization of the results of this study with caution. Future studies should aim to include a more diverse participant pool.

Among the variables of this study, concentration did not yield statistically significant results. However, a comparison between the pretest and posttest outcomes of the experimental and control groups indicated a decline in concentration in the experimental group over time relative to the control group. This underscores the importance of longer-term interventions to determine the impact on concentration more accurately.

## Data Availability

No new data were created or analyzed in this study. Data sharing is not applicable to this study.

